# The role of corticospinal and extrapyramidal pathways in motor impairment after stroke

**DOI:** 10.1101/2022.05.06.22274684

**Authors:** Theresa Paul, Matthew Cieslak, Lukas Hensel, Valerie M. Wiemer, Christian Grefkes, Scott T. Grafton, Gereon R. Fink, Lukas J. Volz

## Abstract

Anisotropy of descending motor pathways has repeatedly been linked to the severity of motor impairment following stroke-related damage to the corticospinal tract (CST). Despite promising findings consistently tying anisotropy of the ipsilesional CST to motor outcome, anisotropy is not yet utilized as a biomarker for motor recovery in clinical practice as a conclusive understanding of degenerative processes in the ipsilesional CST and compensatory roles of other descending motor pathways is hindered by methodological constraints such as estimating anisotropy in voxels with multiple fiber directions, sampling biases, and confounds due to aging-related atrophy. The present study addressed these issues by combining diffusion spectrum imaging (DSI) with a novel compartmentwise analysis approach differentiating voxels with one dominant fiber direction (one-directional voxels) from voxels with multiple fiber directions. Compartmentwise anisotropy for bihemispheric CST and extrapyramidal tracts was compared between chronic stroke patients (N=25), age-matched controls (N=22), and young controls (N=24) and its associations with motor performance of the upper and lower limbs were assessed. Our results provide direct evidence for Wallerian degenration along the entire length of the ipsilesional CST reflected by decreased anisotropy in descending fibers compared to age-matched controls, while aging-related atrophy was observed more ubiquitously across compartments. Anisotropy of descending ipsilesional CST voxels showed a highly robust correlation with various aspects of upper and lower limb motor impairment, highlighting the behavioral relevance of Wallerian degeneration. Moreover, anisotropy measures of two-directional voxels within bihemispheric rubrospinal and reticulospinal tracts were linked to lower limb deficits, while anisotropy of two-directional contralesional rubrospinal voxels explained gross motor performance of the affected hand. Of note, the relevant extrapyramidal structures contained fibers crossing the midline, fibers potentially mitigating output from brain stem nuclei, and fibers transferring signals between the extrapyramidal system and the cerebellum. Thus, specific parts of extrapyramidal pathways seem to compensate for impaired gross arm and leg movements incurred through stroke-related CST lesions, while fine motor control of the paretic hand critically relies on ipsilesional CST integrity. Importantly, our findings suggest that the extrapyramidal system may serve as a compensatory structural reserve independent of post-stroke reorganization of extrapyramidal tracts. In summary, compartment-specific anisotropy of ipsilesional CST and extrapyramidal tracts explained distinct aspects of motor impairment, with both systems representing different pathophysiological mechanisms contributing to motor control post-stroke. Considering both systems in concert may help develop diffusion imaging biomarkers for specific motor functions after stroke.

## Introduction

Stroke-related motor deficits are often caused by damage to the corticospinal tract (CST) and associated white matter (WM) changes are frequently assessed using anisotropy derived from diffusion MRI (dMRI). Studies commonly report a relationship between decreased anisotropy in various parts of the ipsilesional CST and the severity of motor impairment of the upper^1–3^ and lower limbs^4–6^, highlighting anisotropy as a promising biomarker for motor recovery post-stroke.^3^ However, anisotropy measures have yet to find their way into clinical practice as individual predictions of motor impairment and recovery remain challenging for several reasons.

First, most studies use a single measure of motor impairment, hindering a differentiated analysis of various aspects of motor performance. Second, it remains unknown which biological processes may underlie changes in anisotropy or give rise to the correlation between CST anisotropy and motor behavior, rendering conclusive mechanistic interpretations difficult. In line with the notion that the degree of anisotropy reduction reflects the extent of structural damage - often referred to as *microstructural integrity* - lower CST anisotropy coincides with more severe motor impairment.^1^ Given that this correlation can still be observed when calculating anisotropy without including the lesion itself, a commonly held view is that this association is driven by *Wallerian degeneration* of descending fibers, a process that describes how axons passing through the lesion degenerate over time.^7^ However, several methodological limitations hinder a confirmation of this notion as previous dMRI-studies primarily relied on fractional anisotropy (FA) derived from diffusion tensor imaging (DTI). DTI cannot adequately resolve multiple fiber directions within a given voxel,^8^ yielding misleading FA estimates for multidirectional voxels entailing crossing or kissing fibers.^9,10^ Numerous studies have circumvented this issue by focusing the analyses on specific parts of the CST, which contain densely packed, parallel-running descending fibers^11^, therefore thought to reflect the extent of Wallerian degeneration.^12^ Commonly used regions of interest (ROIs) include the pons,^13,14^ the cerebral peduncle (CP),^15,16^ or the posterior limb of the internal capsule (PLIC).^17–19^ Unfortunately, such ROI-based approaches introduce new pitfalls. First, the limited number of voxels within the typically used ROIs aggravates sampling biases.^2^ Moreover, stroke patients usually feature considerable degrees of aging-related WM atrophy^20^ in addition to stroke-induced damage, with certain parts of the brain such as the PLIC being especially prone to aging-related degeneration.^21,22^ Thus, aging-related confounds might bias the quantification of Wallerian degeneration, especially when applying ROI-based approaches. Taken together, this leads to the question whether ipsilesional CST anisotropy is primarily reflective of Wallerian degeneration, which would emphasize its potential as biomarker, or whether aging-related degeneration and methodological limitations may bias the commonly observed correlation with motor performance.

A third aspect that hinders the usage of ipsilesional CST anisotropy as a biomarker is the motor system’s ability to compensate for CST damage via alternate fiber tracts not directly affected by the lesion. Therefore, a patient may not necessarily have to rely on spared fibers of the lesioned CST alone. Specifically, non-crossing fibers of the contralesional CST^23^ or bihemispheric extrapyramidal pathways including the reticulospinal (reticuloST) and rubrospinal tract (rubroST)^1,5^ may compensate for ipsilesional CST damage by supporting basal motor skills via their projections to proximal arm and leg muscles. However, mixed findings hinder a conclusive interpretation of their role in motor control post stroke. While some studies argue that extrapyramidal pathways successfully support gross motor performance, others ascribe a potential maladaptive influence to an upregulated extrapyramidal system, leading to a dysfunctional increase in flexor synergies.^24,25^ While an increase in anisotropy has been conceptualized as an upregulation of the extrapyramidal system caused by structural reorganization post-stroke,^26–28^ other studies could not replicate this finding, even reporting a decrease in anisotropy^29^. Thus, the question arises whether the frequently observed association between extrapyramidal tract anisotropy and motor behavior is a result of structural changes in the extrapyramidal system or whether subjects with a relatively high premorbid level of extrapyramidal anisotropy are better equipped to compensate for CST deficits through reliance on their structural reserve.^30^ Notably, the assessment of anisotropy in extrapyramidal tracts suffers from the same limitations as described above for the CST, rendering the interpretation of anisotropy in the extrapyramidal system challenging.

In sum, a better understanding of the mechanisms underlying degeneration of the CST and behavioral compensation of specific motor functions via alternate motor pathways is crucial to pave the way for the usage of anisotropy as a biomarker in clinical care. To address these issues, we assessed chronic stroke patients using motor tests that differentiate various aspects of motor performance and included age-matched and young controls to discern aging-and stroke-related WM degeneration. Diffusion spectrum imaging (DSI) was employed to better resolve multiple diffusion directions per voxel.^31^ Anisotropy was conceptualized as generalized fractional anisotropy (gFA), which allowed us to quantify the magnitude of multiple intravoxel directions.^32^ Moreover, a compartmentwise analysis approach that differentiates voxels according to their number of trackable directions^10^ (cf. Figure 1) enabled us to separately analyze voxels containing one-directional or multi-directional fibers.

**Figure 1:**
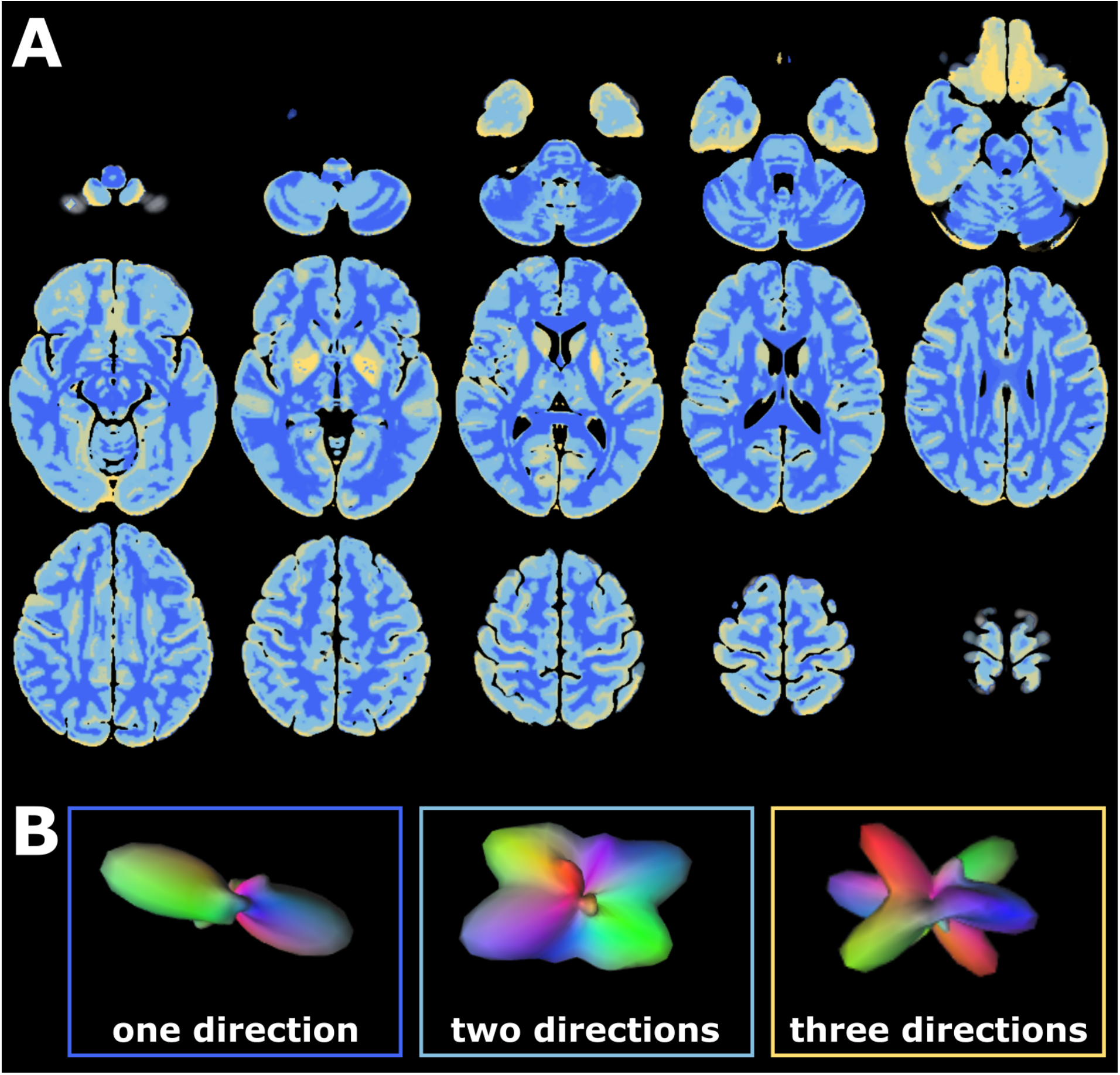
Deterministic brain mask for whole-brain compartmentalization. (A) The mask was created by Volz and colleagues based on 630 subjects from the Human Connectome Project^10^. Each color represents a single compartment containing voxels with a certain number of trackable directions (*dark blue = one direction, light blue = two directions, yellow = three (or more) directions*. Images depicted in (A) were created based on the nifti-file published by Volz et al., 2018.^10^ (B) The number of trackable directions was determined based on the underlying orientation distribution function (ODF) within a given voxel, which can simultaneously depict several diffusion directions. Depending on the number of peaks exceeding a certain threshold, each voxel was assigned to a specific compartment representing the number of trackable intravoxel directions. Exemplary ODFs with different numbers of peaks are shown in (B).

By focusing on voxels containing only one dominant fiber direction, we assessed anisotropy of descending fibers along the length of the entire ipsilesional CST, overcoming the limitations of ROI-based approaches. Assuming that the correlation with motor impairment was genuinely driven by Wallerian degeneration, we hypothesized a positive correlation between motor performance and anisotropy mainly driven by one-directional CST voxels. Given the seminal role of the CST, we expected this correlation to emerge for both basal and complex motor skills of the upper and lower limbs. In line with the hypothesis that other descending motor tracts can partially compensate stroke-related deficits in gross motor control,^4,26,28,29^ we expected anisotropy within contralesional CST and extrapyramidal tracts to be linked to basal arm and leg functions. We further hypothesized a combination of alternate motor pathways and ipsilesional CST to explain more behavioral variance than the latter alone as motor performance ultimately depends on both degenerative and compensatory processes. Finally, if compensation was the result of an upregulation of the extrapyramidal system caused by structural reorganization, we would expect an increase in anisotropy in patients compared to age-matched controls, while no group difference would imply the reliance on a structural reserve instead.

## Materials and methods

### Subjects

25 chronic stroke patients (20 male, 5 female, mean age=66.68, std=11.25) formerly hospitalized at the University Hospital Cologne, Department of Neurology, 22 age-matched controls (16 male, 6 female, mean age=67.05, std=6.59), and 24 young control subjects recruited at the University of California, Santa Barbara (8 male, 16 female, mean age=22.29, std=3.66) were included in this study (see Supplementary Table 1 for patient information, Figure 2 for a lesion overlay). For patients, inclusion criteria were (1) first-ever ischemic stroke before six months or more, (2) initial hand motor deficit, and (3) age 40 to 90 years. Exclusion criteria were (1) any contraindications to MRI, (2) cerebral hemorrhage, (3) bihemispheric infarcts, (4) reinfarction or any other neurological disease, as well as (5) persistence of severe aphasia or neglect. All participants provided informed consent prior to participation. The local ethics committee approved the study carried out under the declaration of Helsinki.

**Figure 2:**
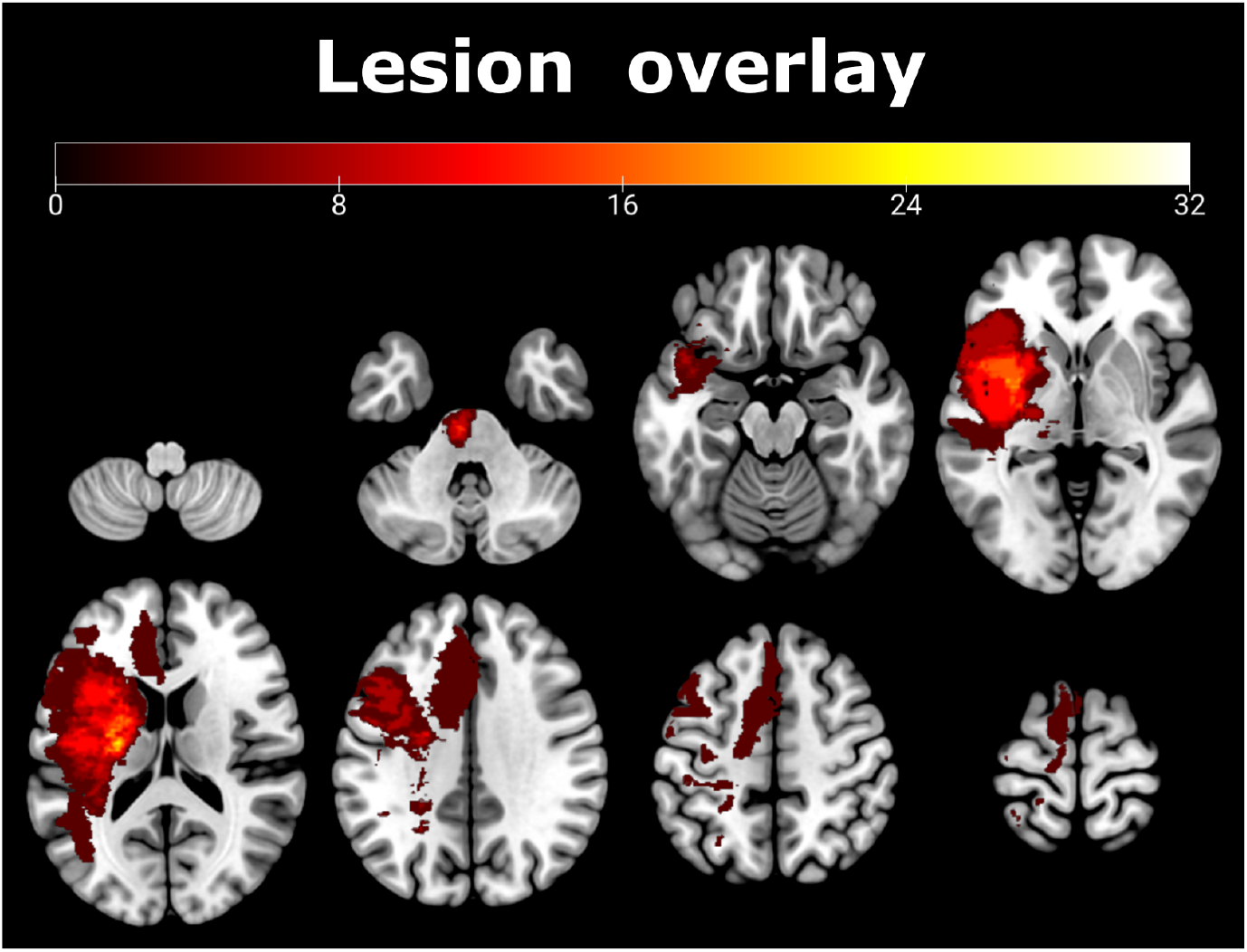
Lesion overlay showing affected voxels across all 25 patients in the left hemisphere (indicated as a percentage). Please note that lesions affecting the right hemisphere were flipped to the left for ease of comparison. The maximum overlap was observed within the internal capsule.

### Motor tests

The motricity index (MI) was assessed in each patient to determine basal motor performance focusing on individual joints of the upper and lower limbs.^33^ For motor skills representing activities of daily living, we used the Action Research Arm Test (ARAT), including the subscales grasp, grip, pinch, and gross movements.^34^

### MRI acquisition

fMRI data were recorded using a Siemens MAGNETOM Prisma 3 Tesla scanner equipped with a 64-channel head coil (Siemens Medical Solutions, Erlangen, Germany). DSI scans were sampled with a spatial resolution of 1.8*1.8*1.8 mm^3^ with b_max_-value of 5000 s/mm^2^, 128 diffusion directions and 10 additional b0s for post-hoc movement correction (TR = 4300 ms, TE=96 ms, FoV=262 mm). In addition, T1-weighted images (TR=2500 ms, TE=2.22 ms, FoV= 241 mm, 208 axial slices, voxel size=0.94*0.94*0.94 mm^3^) and T2-weighted images (TR=3200 ms, TE=0.566 ms, FoV= 241 mm, 208 axial slices, voxel size= 0.94*0.94*0.94 mm^3^) were acquired.

### MRI preprocessing

Preprocessing was performed using QSIPrep 0.13.0RC1,^35^ based on Nipype 1.6.0.^36^ The exact workflow is described in the printout from QSIPrep: The T1-weighted (T1w) image was corrected for intensity non-uniformity using N4BiasFieldCorrection as implemented in ANTs 2.3.1,^37^ and used as T1w-reference throughout the workflow. The T1w-reference was then skull-stripped with antsBrainExtraction, using OASIS as a target template. Spatial normalization to the ICBM 152 Nonlinear Asymmetrical template version 2009c^38^ was performed through nonlinear registration with antsRegistration (ANTs 2.3.1),^39^ using brain-extracted versions of both T1w volume and template. Brain tissue segmentation of cerebrospinal fluid (CSF), WM, and gray matter (GM) was performed on the brain-extracted T1w using FAST as implemented in FSL.^40^

For diffusion data preprocessing, any images with a b-value less than 100 s/mm^2^ were treated as a b=0 image. MP-PCA denoising as implemented in Mrtrix3’s dwidenoise^41^ was applied with a 5-voxel window. B1 field inhomogeneity was corrected using dwibiascorrect from Mrtrix3 with the N4 algorithm.^37^ After B1 bias correction, the mean intensity of the DWI series was adjusted so all the mean intensity of the b=0 images matched across each separate DWI scanning sequence. Motion correction was performed using 3dSHORE as implemented in QSIPrep.^42^ The DWI time series were resampled to ACPC, generating a preprocessed DWI run in ACPC space with 1.8 mm isotropic voxels.

### DSI-reconstruction

Diffusion orientation distribution functions (ODFs) were reconstructed using generalized q-sampling imaging (GQI)^43^ with a ratio of mean diffusion distance of 1.25. Next, individual gFA-maps were created and normalized to the MNI-standard space using ANTs.

### Lesion masking and whole-brain compartmentalization

For patient data, individual lesion masks were drawn on T2-weighted images using MRIcron (www.sph.sc.edu/comd/rorden/Mricron) and applied to patients’ gFA-maps, thereby excluding direct lesion effects on gFA from further analyses to focus on secondary degeneration (cf. Figure 3A). Moreover, individual WM masks derived from QSIPrep were applied to each subject’s gFA-map, excluding non-WM voxels from further analyses. A deterministic mask denoting the number of trackable directions per voxel (constructed using n=630 HCP-datasets)^10^ was used to compartmentalize whole-brain gFA-maps (cf. Figure 1A). Compartmentwise mean gFA-values ((a) all voxels, voxels with (b) one, (c) two, and (d) three fiber directions) were extracted from the motor tracts of interest (CST, rubroST, reticuloST) as defined in the HCP tractography atlas^44^ (cf. Figure 3B). To focus on descending fibers and exclude the widespread cortical inputs to the reticuloST, the reticuloST-mask was trimmed, retaining only the part of the tract below z = -7. In order to avoid a sampling bias induced by different numbers of voxels in left-and right-hemispheric masks, we constructed symmetric masks by flipping left-hemispheric masks along the mid-sagittal plane and applied the resulting masks to the right hemisphere.

**Figure 3:**
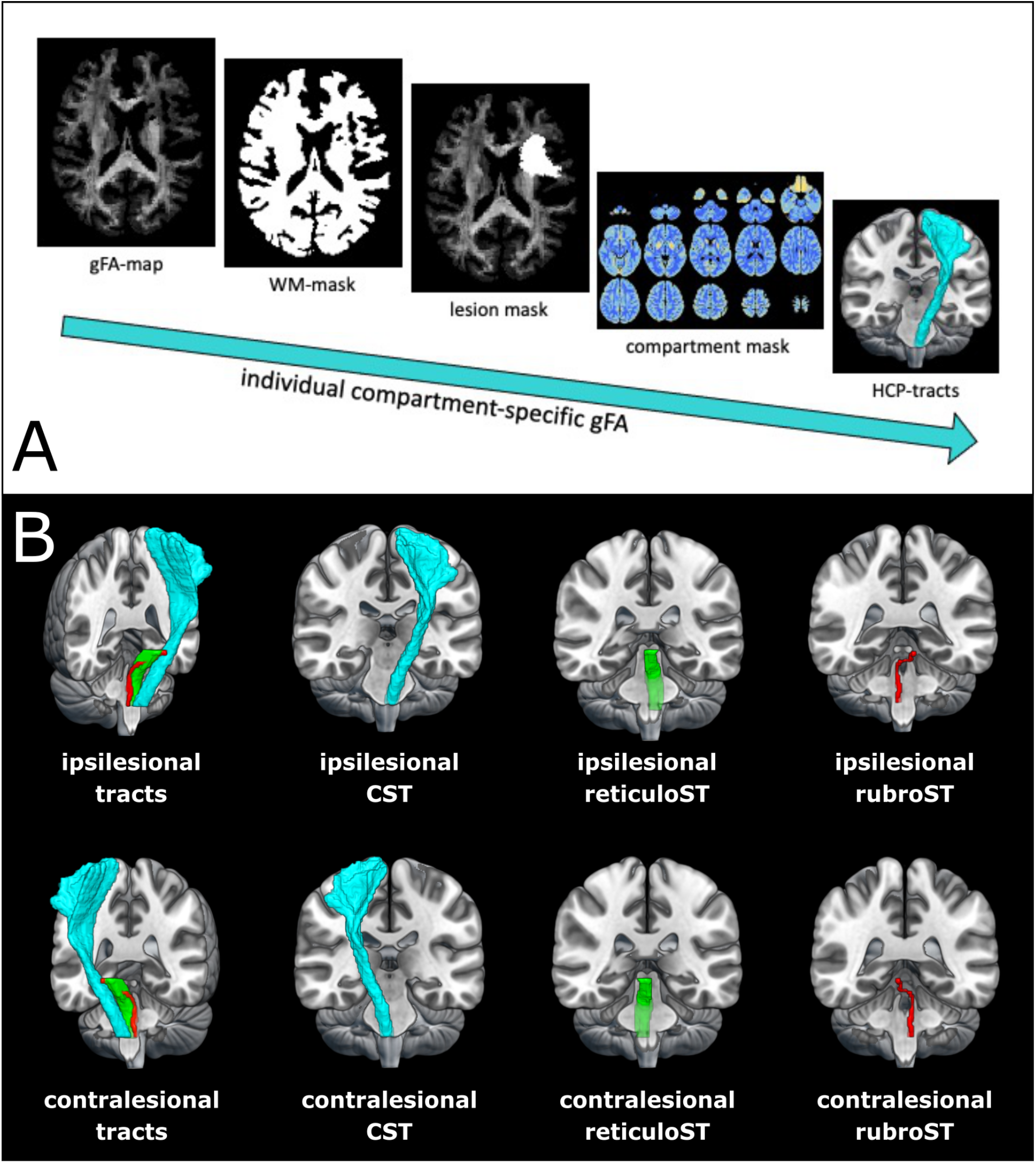
Extraction of compartment-specific tractwise gFA. (A) Workflow to obtain compartment-specific gFA-values per tract. First, voxels containing WM were extracted from each subject’s normalized gFA-map using individual WM tissue classifications as derived from QSIPrep. For patient data, lesion masks were applied to focus the analysis on secondary degeneration processes rather than the assessment of anisotropy within the lesion. A deterministic compartment mask was applied to categorize all voxels according to the number of trackable directions.^10^ Mean gFA was extracted for descending motor tracts as defined by the HCP-tractography atlas.^44^ (B) Motor tracts derived from the HCP tractography atlas^44^ used for gFA-extraction. Note that “ipsilesional” is defined by the origin of the tract superior to its decussation. Symmetric masks were used to ensure an equal number of voxels in each hemisphere, reducing potential sampling biases. *blue = corticospinal tract, green = reticulospinal tract, red = rubrospinal tract*

### Differentiating stroke and aging-related effects

To differentiate between aging- and stroke-related effects, we compared the three subject groups across different compartments for the left and right CST. We computed a repeated-measures ANOVA with the between factor group (levels: patients, age-matched controls, young controls) and the within factor compartment (levels: all (all voxels), one (one-directional voxels), two (two-directional voxels) for the left and right CST. A Greenhouse-Geisser correction was applied where appropriate. Post-hoc pairwise t-tests were used to test for the following differences: (1) compartment all: patients vs. old controls, (2) compartment one: patients vs. old controls, (3) compartment two: patients vs. old controls, (4) compartment all: young vs. old controls, (5) compartment one: young vs. old controls, (6) compartment two: young vs. old controls. Results of post-hoc tests were FDR-corrected for the number of comparisons.

### Probing for extrapyramidal anisotropy differences

By comparing patients and age-matched controls with respect to mean gFA derived from extrapyramidal tracts, we probed for a potential reorganization of the extrapyramidal system. We computed a repeated-measures ANOVA with the between factor group (levels: patients, age-matched controls) and the within factor compartment (levels: all (all voxels), one (one-directional voxels), two (two-directional voxels). A Greenhouse-Geisser correction was applied where appropriate.

### Explaining motor impairment: Ipsilesional CST

In order to probe for a possible relationship with behavior, we computed simple linear regressions with mean gFA derived from the ipsilesional CST as predictor and ARAT, MI-arm, or MI-leg as the outcome variable. Next, we repeated these analyses using mean gFA derived from one- or two-directional voxels as predictors. Moreover, according analyses were performed using the compartment-specific asymmetry index as predictor, which was determined as asymmetry = (mean gFA(unaff CST) – mean gFA(aff CST)) / (mean gFA(unaff CST) + mean gFA(aff CST)).^45^ This step was implemented to improve the signal-to-noise ratio by accounting for the individual aging-related level of atrophy.^16,45^ As previous research has shown that the microstructural integrity of the CST and motor impairment exhibit a stronger relationship in patients with persisting motor deficits,^4,46^ we repeated the analyses for a non-fully recovered subgroup as defined by an ARAT score < 57. For each step, all p-values were FDR-corrected for the number of comparisons.

### Explaining motor impairment: Contralesional CST and extrapyramidal pathways

The role of alternative motor pathways in motor function after stroke was assessed through linear regression analyses. To test for a relationship with basal motor performance, five linear regression models were computed with the MI-arm score as the outcome variable and contralesional CST, ipsilesional reticuloST, contralesional reticuloST, ipsilesional rubroST or contralesional rubroST as the predictor variable. The resulting p-values were FDR-corrected for the number of comparisons. The same procedure was repeated for the ARAT score and the MI-leg score as outcome variable to probe for a potential relationship with the performance of activities of daily living or lower limb performance, respectively. Next, we tested for shared variance between alternative motor pathways and the ipsilesional CST by entering mean gFA-values derived from extrapyramidal tracts that correlated with motor performance into the regression model containing gFA derived from one-directional ipsilesional CST voxels. An overlay of all four extrapyramidal tracts was created to test whether compartment two of the four tracts captured a high number of identical voxels.

## Data availability statement

Data are available from the corresponding author upon reasonable request.

## Results

### Aging-versus stroke-related CST-anisotropy

For the ipsilesional CST, we found a significant main effect of group (F(2, 68)=32.28, p<.001, generalized η^2^=0.44) and compartment (F(1.05, 71.56)=2178.42, p<.001, generalized η^2^= 0.84) as well as an interaction between group and compartment (F(2.10, 71.56)=19.77, p<.001, generalized η^2^=0.09). Post-hoc independent sample t-tests showed that patients featured reduced gFA-values within the ipsilesional CST compared to age-matched controls when considering all voxels within the CST-mask (t(45)=-2.65, p=.013, FDR-corrected). This difference was attributable to voxels containing only one fiber direction (t(45)=-3.66, p=.001, FDR-corrected), while two-directional voxels showed no difference (t(45)=0.41, p=.684, FDR-corrected; cf. Figure 4A). At the same time, young and old controls differed with respect to all levels of the factor compartment (all p<.001, FDR-corrected), indicating that the age-related difference could be objectified in all compartments, affecting both one-and two-directional voxels. For the contralesional CST, we found a main effect of group (F(2,68)=19.24, p<.001, generalized η^2^=0.34) and a main effect of compartment (F(1.06, 71.80)=3756.75, p<.001, generalized η^2^=0.86). There was no interaction between group and compartment (F(2.11, 71.80)=2.41, p=.094). While patients and age-matched controls did not differ with respect to mean gFA in any compartment (p>.1), young and elderly control subjects differed across all compartments (p<.01, FDR-corrected; cf. Figure 4B). Thus, while stroke-related changes were specific to the ipsilesional CST, aging-related degeneration in anisotropy affected the CST in both hemispheres.

**Figure 4:**
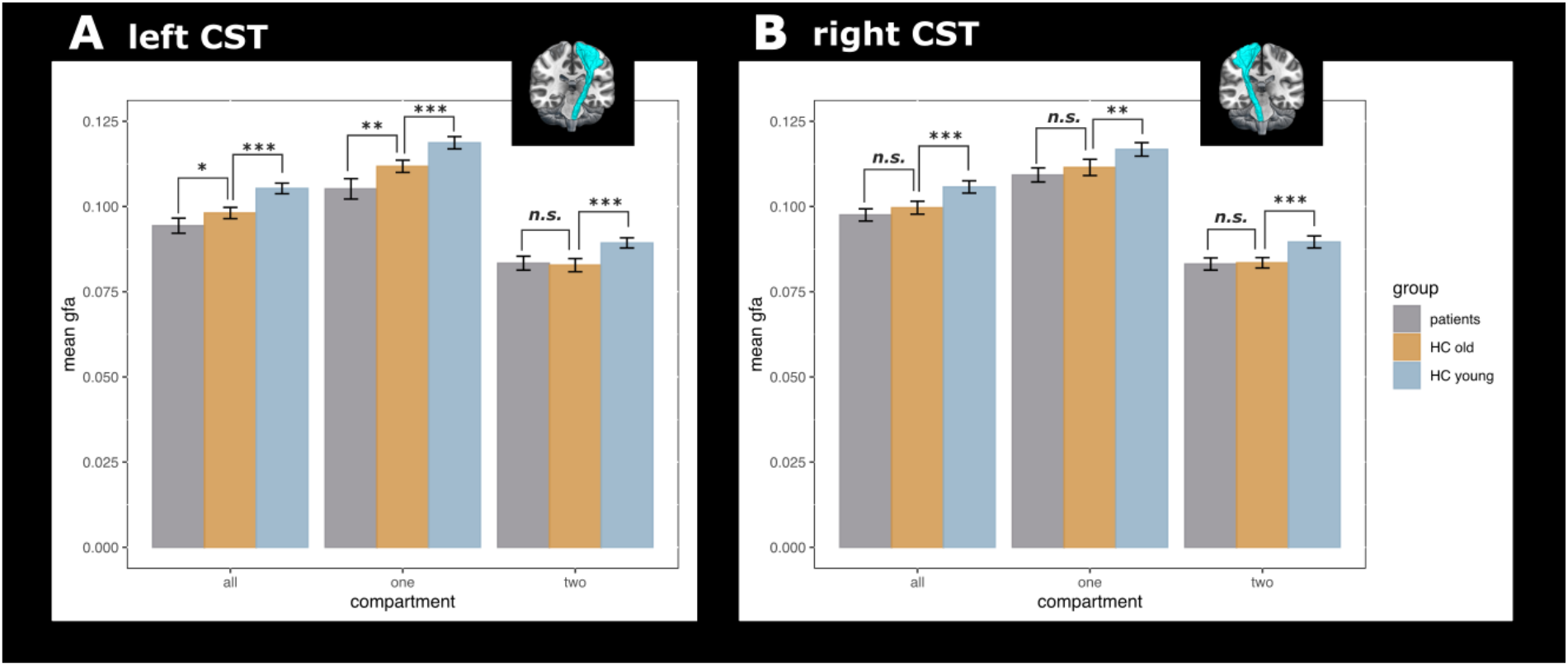
Group differences per compartment. (A) Left (ipsilesional) CST: While young and old controls differed across all compartments (all: all voxels, one: one-directional voxels, two: two-directional voxels), patients and age-matched controls differed for compartment one but not for compartment two. Thus, aging-related changes occurred across the entire left CST entailing both one- and two-directional voxels, while stroke-related decreases in gFA were driven by one-directional voxels. (B) Right (contralesional) CST: In line with findings for the left CST, young and old controls differed across all compartments, yet there was no difference between patients and age-matched controls. Significance thresholds: *** p < .001, ** p < .01, * p < .05. Error bars represent two standard errors.

### Anisotropy in the extrapyramidal system

For the extrapyramidal system entailing bihemispheric reticuloST and rubroST, there was a main effect of compartment (F(1.01, 45.29)=656.22, p<.001, generalized η^2^=0.38) but no main effect of group (F(1,45)=0.01, p=.913) or interaction between group and compartment (F(1.01,45.29)= 0.78, p=.381), indicating that patients and age-matched controls did not differ with respect to extrapyramidal anisotropy.

### Ipsilesional CST degeneration explains motor impairment

Mean gFA-values derived from all voxels of the ipsilesional CST significantly explained upper limb impairment as measured by the ARAT (R^2^=34.61, p=.006, FDR-corrected) or MI-arm (R^2^=28.9%, p=.008, FDR-corrected; cf. Figure 5). The regression model containing MI-leg as the outcome variable showed a trend towards significance (R^2^=15.25%, p=.054, FDR-corrected). Repeating the analyses with only one-directional voxels showed that the results were indeed driven by descending fibers (ARAT: R^2^=30.94%, p=.006; MI-arm: R^2^=31.94%, p=.010; MI-leg: R^2^=17.71%, p=.036, all FDR-corrected). Regression models using mean gFA derived from compartment two did not reach significance (all p>.2, FDR-corrected). Using the asymmetry index based on the entire CST as the predictor yielded a slightly higher percentage of explained variance for the ARAT (R^2^=36.57%, p=.004, FDR-corrected) but not for MI-arm (R^2^=28.10%, p=.010, FDR-corrected) or MI-leg (R^2^=11.21%, p=.102, FDR-corrected). Increased explained variance was also observed when computing the asymmetry index for one-directional voxels (ARAT: R^2^=34.11%, p=.007; MI-arm: R^2^=31.78%, p=.005; MI-leg: R^2^=16.01%, p=.048, all FDR-corrected).

**Figure 5:**
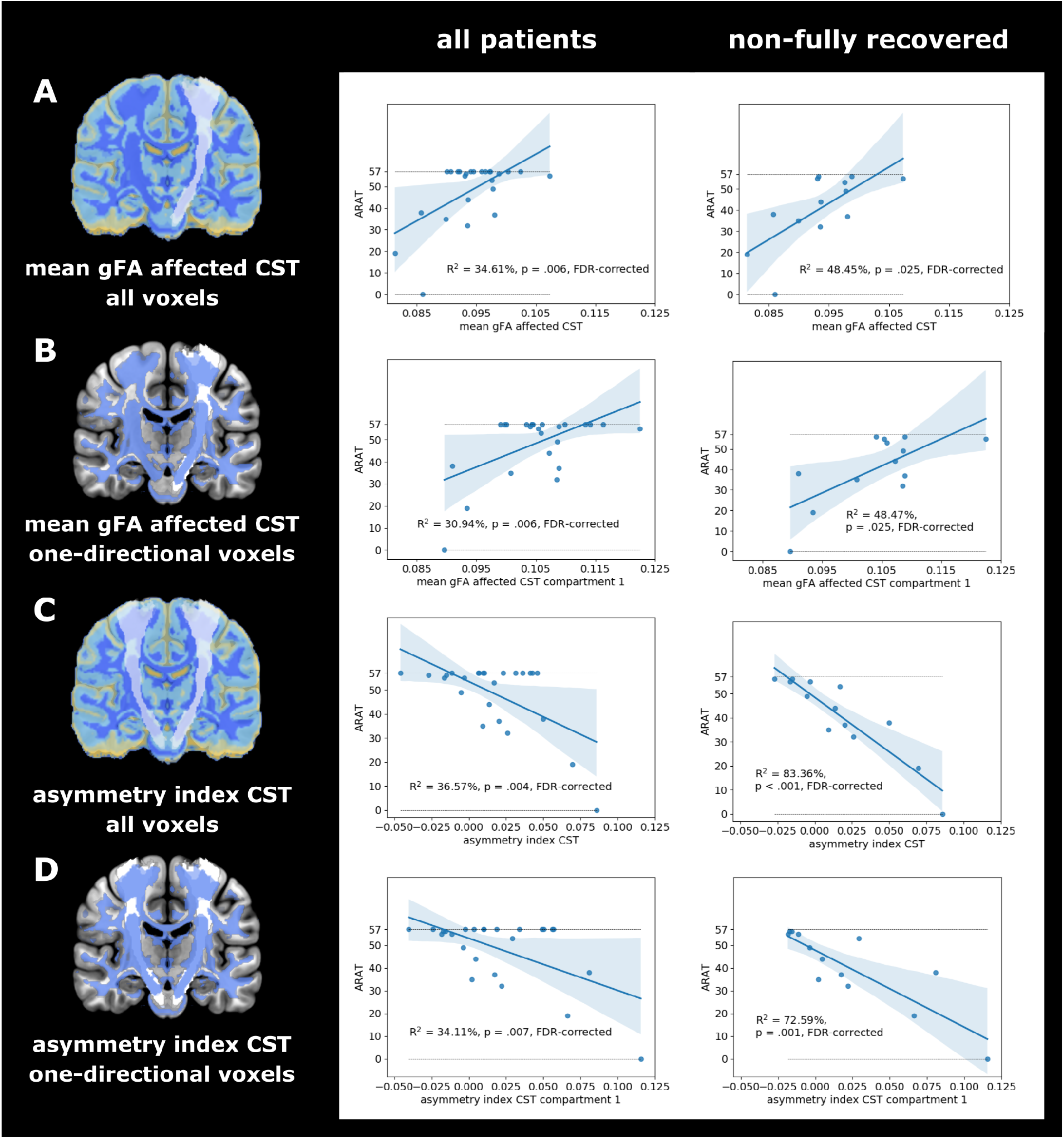
Regression analyses for the association between different gFA-based CST measures and ARAT motor scores. (A) Mean gFA-values within the ipsilesional CST explained motor impairment for the entire cohort and to an even higher degree for the non-fully recovered subgroup. (B) By obtaining the mean gFA-value from one-directional voxels within the ipsilesional CST, it becomes evident that the association between anisotropy and motor performance was driven by descending fibers. Note that the mean taken from two-directional voxels did not significantly explain motor performance (p>.2, FDR-corrected). (C) To improve the signal-to-noise ratio, we calculated the asymmetry index that accounts for the subject-specific level of atrophy by considering both the affected and unaffected CST. Across all CST-voxels, we observed a significant association with the ARAT score for the entire sample and the non-fully recovered subgroup. (D) Entering the asymmetry index from one-directional voxels as the predictor into the regression models yielded a similar ratio of explained variance as in (C).

When exclusively considering patients with persisting motor deficits and repeating the analyses for this non-fully recovered subgroup, the ratio of explained variance increased for all models, when using mean gFA across all voxels (ARAT: R^2^=48.45%, p=.025; MI-arm: R^2^=40.77%, p=.028; MI-leg: R^2^=29.12%, p=.057, all FDR-corrected) or gFA extracted from one-directional voxels (ARAT: R^2^=48.47%, p=.025; MI-arm: R^2^=46.22%, p=.016; MI-leg: R^2^=35.11%, p=.033, all FDR-corrected), as well as the asymmetry index derived from all voxels (ARAT: R^2^=83.36%, p<.001 MI-arm: R^2^=63.08%, p=.002; MI-leg: R^2^=50.88%, p=.006, all FDR-corrected) or one-directional voxels (ARAT: R^2^=72.59%, p=.001; MI-arm: R^2^=63.08%, p=.002; MI-leg: R^2^=52.47%, p=.005, all FDR-corrected).

### Extrapyramidal pathways explain motor impairment

While neither the contralesional CST nor any of the extrapyramidal tracts explained variance in ARAT scores in any compartment (p>.7, FDR-corrected), the rubroST descending from the contralesional hemisphere showed a significant relationship with the MI-arm score for two-directional voxels (R^2^=25.94%, p=.047, FDR-corrected) and a trend towards significance when using all voxels (R^2^=22.18, p=.087, FDR-corrected). For the MI-leg score, all extrapyramidal tracts explained variance in motor performance when using two-directional voxels (all p<.05), but not the contralesional CST (cf. Table 1 and Figure 6). Notably, the two-directional voxels of the four extrapyramidal tracts barely overlapped, with only seven voxels being included in all four masks, indicating that the explained variance observed for the different tracts was not driven by the same set of overlapping voxels (cf. Figure 6F).

**Table 1:** Linear regression results of the motricity index leg score for alternative motor output pathways using mean gFA derived from two-directional voxels.

| predictor | R<sup>2</sup> | p (FDR) |
| --- | --- | --- |
| contralesional CST | 1.11% | 0.616 |
| ipsilesional reticuloST | 19.14 % | 0.036 |
| contralesional reticuloST | 21.85 % | 0.046 |
| ipsilesional rubroST | 21.77 % | 0.031 |
| contralesional rubroST | 27.37% | 0.036 |

**Figure 6:**
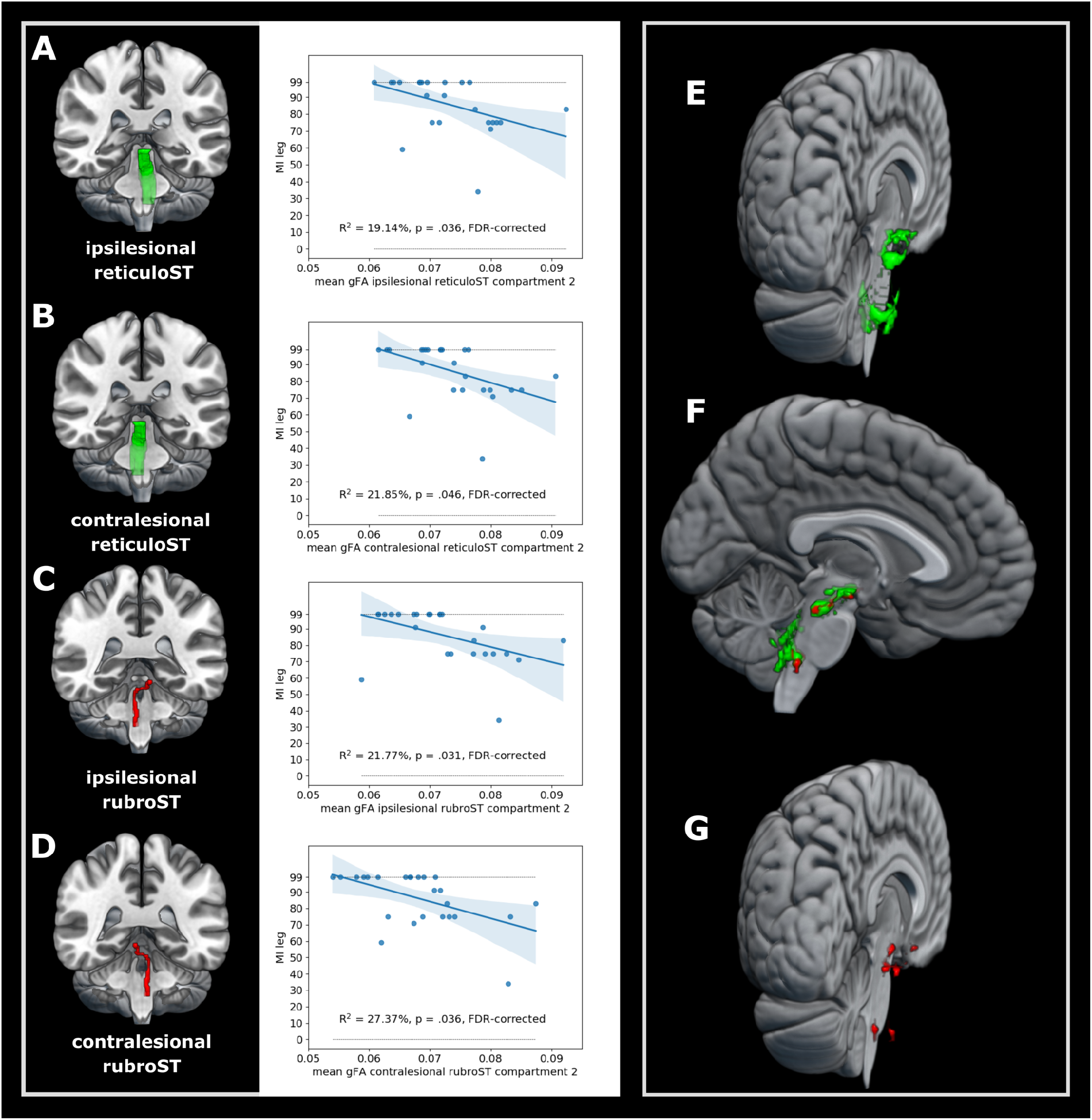
Explained variance by mean gFA derived from two-directional voxels of extrapyramidal pathways for the motricity index lower extremity scores. Note that there was a robust negative relationship between leg impairment and gFA derived from two-directional voxels within the (A) ipsilesional reticuloST, (B) contralesional reticuloST, (C) ipsilesional rubroST, and (D) contralesional rubroST. Of note, compartment two voxels of the reticuloST (E) and rubroST (G) did not contain many overlapping voxels (F).

To assess whether the extrapyramidal system’s anisotropy may reflect a compensatory mechanism, i.e., may hold additional information on motor outcome exceeding that of the ipsilesional CST, we tested whether the extrapyramidal system explained additional behavioral variance when combined with the ipsilesional CST. Adding mean gFA derived from compartment two of any of the extrapyramidal tracts as an additional variable to anisotropy derived from one-directional ipsilesional CST voxels into a regression model significantly increased the amount of explained variance for the MI-leg score, underlining the independence of extrapyramidal tracts from the ipsilesional CST (all R^2^-values>38%, p<.01, cf. Supplementary Table 2). Concerning the shared variance between ipsilesional CST and extrapyramidal system in the prediction of gross upper limb performance, the combination of mean gFA derived from compartment one of the ipsilesional CST and compartment two of the contralesional rubroST led to a significantly higher ratio of explained variance than any of the two tracts alone (R^2^=51.02%, p<.001).

## Discussion

Using DSI data combined with a compartmentwise analysis approach that differentiates the number of fiber directions per voxel, we found that decreased anisotropy following stroke primarily affected one-directional voxels of the ipsilesional CST while aging-related degeneration was observed across all directional compartments. In line with numerous previous findings, ipsilesional CST anisotropy explained motor performance across many functional domains, which entirely relied on one-directional voxels representing descending fibers. Thus, our data provide direct evidence for Wallerian degeneration occurring throughout the entire ipsilesional CST and underline the seminal pathophysiological role of Wallerian degeneration for various aspects of motor function. However, the ipsilesional CST cannot be considered the sole descending motor pathway involved in motor control post-stroke: anisotropy of extrapyramidal tracts was associated with specific aspects of motor impairment, highlighting function-specific compensatory roles of distinct pathways. While the contralesional rubroST was indicative of gross motor control of the arm, bihemispheric rubroST and reticuloST were related to lower limb motor function. Importantly, the relationship between extrapyramidal tracts and motor performance only emerged when focusing on voxels containing two fiber directions, which may explain contradictory findings of previous studies. Of note, stroke patients did not differ from age-matched controls regarding anisotropy in extrapyramidal tracts, suggesting that functional compensation through extrapyramidal pathways does not rely on reorganization of these tracts but rather reflects an aspect of the structural reserve of the motor system as discussed below.

### Structural CST alterations in stroke and aging

Using a compartmentwise approach, we aimed to disentangle the drivers of anisotropy changes in chronic stroke patients to further elucidate the potential of anisotropy in descending motor tracts to explain motor control after stroke. In line with our hypotheses, the expected decline in anisotropy in the ipsilesional CST compared to age-matched controls was limited to voxels containing one-directional, i.e., descending fibers, and could not be observed in the contralesional CST (cf. Figure 4). Conversely, age-related differences between the young and old control group manifested themselves in bilateral CSTs regardless of the number of intravoxel directions, which nicely fits the notion that aging represents a global phenomenon affecting more than just descending fiber tracts.^20^ Thus, the compartmentwise analysis approach allowed us to differentiate aging- and stroke-related anisotropy changes. Of note, decreased ipsilesional CST anisotropy in stroke patients compared to age-matched controls was only evident in one-directional voxels, highlighting Wallerian degeneration as the main driver of anisotropy changes in descending ipsilesional CST fibers. The pivotal influence of Wallerian degeneration on anisotropy changes post-stroke clarifies why previous research has frequently linked the integrity of ipsilesional CST fibers to motor impairment. For instance, Schaechter and colleagues investigated differences between chronic stroke patients and healthy controls by comparing FA curves extracted along the ipsilesional CST starting from the precentral gyrus down to the CP.^23^ Their results indicate that poorly recovered stroke patients feature significantly decreased FA values in the ipsilesional CST between the height of the PLIC and CP. Parts of this CST section have repeatedly been used as ROIs to investigate stroke-related WM abnormalities and their association with motor impairment. As Pierpaoli and colleagues point out, this particular stretch of the CST is characterized by densely packed descending fibers without any significant association tracts passing through.^11^ In other words, it mainly consists of descending fibers which we captured as one-directional voxels in our analyses.

Interestingly, our results suggest that focusing on descending fibers along the entire CST might increase the sensitivity for certain aspects of motor performance: When using all CST voxels, mean gFA significantly explained motor impairment of the upper but not the lower limb. However, lower limb deficits could also be explained when exclusively including one-directional voxels. Moreover, the significant association between gFA in the ipsilesional CST and motor impairment of the upper limb was entirely driven by one-directional voxels: While mean gFA derived from one-directional voxels accounted for almost the same amount of explained variance as all CST-voxels combined, two-directional voxels did not show any association with motor impairment. Of note, these effects were intensified for the non-fully recovered subgroup, which is in line with previous studies reporting more severe WM changes and better predictions of motor performance in patients with more pronounced motor deficits.^4,46^ The ratio of explained variance was even higher when using the asymmetry index and thereby accounting for a patient’s individual level of aging-related atrophy.

Importantly, the present findings do not only underline the applicability of a DSI-based compartmentwise approach, but also offer a possible solution to the problem of arbitrary ROI selection since focusing on descending fibers allowed us to include the entire length of the CST into our analysis. Moreover, using one-directional voxels from the entire length of the ipsilesional CST may also help to reduce aging-related confounds commonly introduced when focusing on regions heavily affected by aging-related atrophy such as the PLIC.^21,22^ In summary, anisotropy of one-directional ipsilesional CST fibers primarily reflects Wallerian degeneration of descending motor fibers which accounted for a large amount of variance in motor impairment across various domains of motor control. However, our current results also highlight that Wallerian degeneration of the ipsilesional CST should not be regarded as the only factor contributing to motor control after stroke, given the compensatory potential of other descending motor pathways.

### Compensatory role of the extrapyramidal system

The role of the extrapyramidal system in motor recovery following stroke is a subject of ongoing debate, inspired by several studies reporting associations between motor impairment and extrapyramidal tract anisotropy. While some authors argue that increased reliance on the extrapyramidal system may help patients to recover successfully,^4,29^ others interpret their findings as maladaptive reorganization.^24,27,28,47^ These opposing interpretations are largely driven by the direction of the observed correlations between anisotropy and motor behavior: positive correlations linking higher anisotropy to better motor performance are often construed as beneficial compensation, whereas negative correlations linking higher anisotropy to worse motor performance are commonly interpreted as maladaptive overcompensation caused by reorganization processes of the extrapyramidal tracts. A commonly held view is that the overcompensation may stem from an overactivation of extrapyramidal pathways resulting in increased flexor synergies, hindering the control of individual movements.^24,25^ Here, we exclusively observed negative associations between extrapyramidal anisotropy and motor performance of the arm and leg (cf. Figure 6). Therefore, one may interpret our current findings as a maladaptive over-compensation by the extrapyramidal system. However, motor impairment across patients was exclusively explained by two-directional voxels which renders the interpretation more difficult. Two-directional voxels are characterized by two dominant directions which need to change to a different extent for gFA to either de- or increase. Depending on the ratio of the two directions, even an underlying decrease in one direction could lead to an overall increase in gFA. For instance, if only the non-dominant direction in a two-directional voxel decreases while the dominant direction remains constant, the gFA-value will increase. Following this logic, gFA-values will not change at all when both fiber directions within a two-directional voxel change to the same extent. Thus, a higher or lower degree of anisotropy in two-directional voxels should not be mistaken for higher or lower microstructural integrity. In other words, higher or lower degrees of anisotropy in two-directional voxels cannot be functionally interpreted in a straightforward way, which may well account for previous contradictory findings. Therefore, future studies should focus on changes at the subvoxel level to further our understanding of the underlying pathophysiological mechanisms.

From a functional perspective, extrapyramidal pathways are thought to support gross motor function via their projections to proximal muscles of the arm and leg. In particular, the basal motor control by extrapyramidal tracts may stem from their ability to directly code for strength of muscle activation as recently observed for the reticular formation in monkeys.^48^ Our current results are well in line with this notion, as reflected by their associations with MI scores of the upper and lower limb but not for the ARAT. For the ARAT, higher scores can be achieved through compensatory strategies applied in daily life, which requires higher degrees of motor control and more complex motor skills.^49^ The MI, on the other hand, assesses each joint individually. In other words, it relies on muscular strength and therefore reflects more basal demands on the motor system.^50,51^ Given the observed relationship between anisotropy of all four extrapyramidal tracts and the patients’ ability to move individual joints as assessed using the MI, our current findings underline the potential involvement of extrapyramidal pathways in the recovery of gross motor function after stroke. This notion is supported by the fact that extrapyramidal pathways seemed to be independent of the ipsilesional CST as indicated by the increase in explained variance when combining CST and extrapyramidal anisotropy in a regression model explaining MI scores. These observations perfectly match previous reports indicating an additional explanation of variance in motor function by anisotropy in bilateral rubroST independent of ipsilesional CST.^29^ Thus, these findings are in line with the notion that both pyramidal and extrapyramidal tracts contribute independently to the execution and control of gross motor function. To further elucidate the mechanistical role of extrapyramidal tracts in motor control post-stroke, a seminal question lies in whether extrapyramidal tracts undergo stroke-induced reorganization or whether compensation may be determined by the premorbid level of descending motor output. Considering that we did not find a group difference between stroke patients and age-matched controls, a compensatory upregulation through structural changes of the extrapyramidal tracts seems less likely. Conversely, extrapyramidal anisotropy may rather indicate a structural reserve within those tracts the motor system may capitalize on to compensate for stroke-induced damage.^30^

### Anatomical foundations of extrapyramidal compensation

Having established a link between motor function and extrapyramidal integrity of two-directional voxels, the question arises which anatomical parts of the reticuloST and rubroST drove this observation. The reticuloST receives its cortical inputs from the primary motor cortex, as well as the premotor and supplementary motor areas^52^ and descends mostly ipsilaterally from the medial pontine and medullary reticular formation^53^. The rubroST originates from the red nucleus at the level of the mesencephalon and receives inputs from an array of cortical areas and the cerebellum. It decussates at the level of the red nucleus and descends alongside the lateral CST.^54^ While most fibers within the rubroST decussate, most of the reticuloST descends without crossing. Therefore, it seems striking that tracts from both hemispheres showed a similar relationship with motor performance. A potential explanation for this observation might derive from the limited number of crossing fibers in both tracts.^5,55^ Considering the anatomical proximity of these crossing fibers and the relatively high overlap in explained variance in motor performance of all four tracts, one might assume that identical voxels were included in both tracts due to tracking inaccuracies or sampling biases. However, computing the overlap between all four masks yielded a negligible number of shared voxels.

When visualizing compartment two voxels (cf. Figure 6E-G), clusters emerged at three different levels. Voxels located at the most superior level may potentially constitute input and output fibers from various nuclei. For example, anisotropy surrounding the red nucleus has been shown to correlate with motor impairment,^4,26,28^ supporting the notion that output properties of the red nucleus may contribute to motor control after stroke. Moreover, units in the pontine reticular formation of the cat discharge during motor activity even when deprived of any other stimulus inputs,^56^ which highlights the involvement of the pontine reticular formation and the descending reticuloST for the generation of motor output. Second, various clusters can be seen close to the midline, where both rubroST and the reticuloST cross over to the other hemisphere. Whether the integrity of these crossings is vital for motor performance after stroke remains an interesting question for future research. Third, at the lowest level, a cluster of two-directional voxels may include fibers from cerebellar structures. Cortico-cerebellar pathways have already been shown to relate to residual motor function after stroke^57,58^ and it might be possible that the observed relationship with motor impairment was partially driven by fibers projecting to or from the cerebellum. In summary, our methodological approach helped to identify specific parts of the extrapyramidal system contributing to compensation of gross arm and leg movements after stroke, primarily comprising fibers crossing the midline, fibers potentially mitigating output from brain stem nuclei (such as the red nucleus), or interactions across different parts of the extrapyramidal system and cerebellum.

### Limitations and future directions

The focus on chronic stroke patients in the present study allowed us to draw conclusions regarding the effects of secondary degenerative processes and to assess the reorganized motor system. Further studies are warranted to quantify anisotropy changes in descending tracts longitudinally starting in the acute phase after stroke. As our findings suggest that compensatory processes in the extrapyramidal system rely on a patient’s structural reserve rather than structural changes within the extrapyramidal tracts, estimating the propensity of these tracts to compensate for lost functionality based on anisotropy measures might already be possible in the acute phase post-stroke. Moreover, the present study focused on structural alterations in descending tracts underlining their relationship to motor impairment. Given that some compensatory mechanisms may also operate through cortico-cortical interactions, extending the current data by assessing structural connectivity between different cortical motor areas may result in a more comprehensive picture. Since motor impairment has also been shown to be closely related to functional connectivity between cortical motor regions,^59–64^ future research should try to elucidate the relationship between stroke-related changes in the structural and functional organization of the motor network.

## Conclusion

By disentangling aging from stroke-related effects via compartmentwise analyses, we provide direct evidence for Wallerian degeneration of the ipsilesional descending CST and its seminal role in various aspects of motor control of the upper and lower limbs. Anisotropy of the contralesional rubroST explained gross motor performance of the affected hand, while anisotropy within all extrapyramidal tracts located throughout the brainstem were linked to motor function of the lower limb in chronic stroke patients, supporting the notion of increased reliance on extrapyramidal pathways to support basal motor function after CST damage. Of note, all extrapyramidal tract findings were based on two-directional voxels which can be found in specific anatomical locations throughout the brainstem, potentially mitigating output of brainstem nuclei, signals crossing the midline and cerebellar influences. Since the highest ratio of explained variance was achieved when combining extrapyramidal pathways and ipsilesional CST anisotropy, clinical biomarkers should consider both the degeneration of the ipsilesional CST as well as the compensation via the extrapyramidal system. From a mechanistic perspective, our findings suggest that compensatory processes in the extrapyramidal system reflect an aspect of structural motor reserve rather than reorganization of extrapyramidal tracts. In summary, anisotropy of descending motor pathways seems to be a promising marker for motor impairment post-stroke, especially when divided into compartments based on the number of trackable directions per voxel.

## Data Availability

Data are available from the corresponding author upon reasonable request.

## List of abbreviations

CP: cerebral peduncle
CST: corticospinal tract
dMRI: diffusion magnetic resonance imaging
DSI: diffusion spectrum imaging
DTI: diffusion tensor imaging
FA: fractional anisotropy
gFA: generalized fractional anisotropy
ODF: orientation distribution function
PLIC: posterior limb of the internal capsule
reticuloST: reticulospinal tract
ROI: region of interest
rubroST: rubrospinal tract
WM: white matter

## References

1. Koch P, Schulz R, Hummel FC. Structural connectivity analyses in motor recovery research after stroke. Ann Clin Transl Neurol. 2016;3(3):233–244. doi:10.1002/acn3.278

2. Moura LM, Luccas R, Paiva JPQ de, et al. Diffusion Tensor Imaging Biomarkers to Predict Motor Outcomes in Stroke: A Narrative Review. Front Neurol. 2019;10(MAY):1–17. doi:10.3389/fneur.2019.00445

3. Boyd LA, Hayward KS, Ward NS, et al. Biomarkers of stroke recovery: Consensus-based core recommendations from the Stroke Recovery and Rehabilitation Roundtable. Int J Stroke. 2017;12(5):480–493. doi:10.1177/1747493017714176

4. Peters DM, Fridriksson J, Richardson JD, et al. Upper and Lower Limb Motor Function Correlates with Ipsilesional Corticospinal Tract and Red Nucleus Structural Integrity in Chronic Stroke: A Cross-Sectional, ROI-Based MRI Study. Tambasco N, ed. Behav Neurol. 2021;2021:1–10. doi:10.1155/2021/3010555

5. Cleland BT, Madhavan S. Ipsilateral motor pathways to the lower limb after stroke: Insights and opportunities. J Neurosci Res. 2021;99(6):1565–1578. doi:10.1002/jnr.24822

6. Jayaram G, Stagg CJ, Esser P, Kischka U, Stinear J, Johansen-Berg H. Relationships between functional and structural corticospinal tract integrity and walking post stroke. Clin Neurophysiol. 2012;123(12):2422–2428. doi:10.1016/j.clinph.2012.04.026

7. Conforti L, Gilley J, Coleman MP. Wallerian degeneration: an emerging axon death pathway linking injury and disease. Nat Rev Neurosci. 2014;15(6):394–409. doi:10.1038/nrn3680

8. Basser PJ, Mattiello J, LeBihan D. MR diffusion tensor spectroscopy and imaging. Biophys J. 1994;66(1):259–267. doi:10.1016/S0006-3495(94)80775-1

9. Descoteaux M, Angelino E, Fitzgibbons S, Deriche R. Apparent diffusion coefficients from high angular resolution diffusion imaging: Estimation and applications. Magn Reson Med. 2006;56(2):395–410. doi:10.1002/mrm.20948

10. Volz LJ, Cieslak M, Grafton ST. A probabilistic atlas of fiber crossings for variability reduction of anisotropy measures. Brain Struct Funct. 2018;223(2):635–651. doi:10.1007/s00429-017-1508-x

11. Pierpaoli C, Barnett A, Pajevic S, et al. Water Diffusion Changes in Wallerian Degeneration and Their Dependence on White Matter Architecture. Neuroimage. 2001;13(6):1174–1185. doi:10.1006/nimg.2001.0765

12. Kim B, Fisher BE, Schweighofer N, et al. A comparison of seven different DTI-derived estimates of corticospinal tract structural characteristics in chronic stroke survivors. J Neurosci Methods. 2018;304:66–75. doi:10.1016/j.jneumeth.2018.04.010

13. Puig J, Pedraza S, Blasco G, et al. Wallerian degeneration in the corticospinal tract evaluated by diffusion tensor imaging correlates with motor deficit 30 days after middle cerebral artery ischemic stroke. Am J Neuroradiol. 2010;31(7):1324–1330. doi:10.3174/ajnr.A2038

14. Puig J, Blasco G, Daunis-I-Estadella J, et al. Decreased Corticospinal Tract Fractional Anisotropy Predicts Long-term Motor Outcome After Stroke. Stroke. 2013;44(7):2016–2018. doi:10.1161/STROKEAHA.111.000382

15. Thomalla G, Glauche V, Koch MA, Beaulieu C, Weiller C, Röther J. Diffusion tensor imaging detects early Wallerian degeneration of the pyramidal tract after ischemic stroke. Neuroimage. 2004;22(4):1767–1774. doi:10.1016/j.neuroimage.2004.03.041

16. Doughty C, Wang J, Feng W, Hackney D, Pani E, Schlaug G. Detection and Predictive Value of Fractional Anisotropy Changes of the Corticospinal Tract in the Acute Phase of a Stroke. Stroke. 2016;47(6):1520–1526. doi:10.1161/STROKEAHA.115.012088

17. Borich MR, Mang C, Boyd LA. Both projection and commissural pathways are disrupted in individuals with chronic stroke: investigating microstructural white matter correlates of motor recovery. BMC Neurosci. 2012;13(1):107. doi:10.1186/1471-2202-13-107

18. Puig J, Pedraza S, Blasco G, et al. Acute Damage to the Posterior Limb of the Internal Capsule on Diffusion Tensor Tractography as an Early Imaging Predictor of Motor Outcome after Stroke. Am J Neuroradiol. 2011;32(5):857–863. doi:10.3174/ajnr.A2400

19. Borich MR, Brown KE, Boyd LA. Motor Skill Learning Is Associated With Diffusion Characteristics of White Matter in Individuals With Chronic Stroke. J Neurol Phys Ther. 2014;38(3):151–160. doi:10.1097/NPT.0b013e3182a3d353

20. Kelley S, Plass J, Bender AR, Polk TA. Age-Related Differences in White Matter: Understanding Tensor-Based Results Using Fixel-Based Analysis. Cereb Cortex. 2021;31(8):3881–3898. doi:10.1093/cercor/bhab056

21. Salat DH, Tuch DS, Greve DN, et al. Age-related alterations in white matter microstructure measured by diffusion tensor imaging. Neurobiol Aging. 2005;26(8):1215–1227. doi:10.1016/j.neurobiolaging.2004.09.017

22. Hsu JL, Leemans A, Bai CH, et al. Gender differences and age-related white matter changes of the human brain: A diffusion tensor imaging study. Neuroimage. 2008;39(2):566–577. doi:10.1016/j.neuroimage.2007.09.017

23. Schaechter JD, Perdue KL, Wang R. Structural damage to the corticospinal tract correlates with bilateral sensorimotor cortex reorganization in stroke patients. Neuroimage. 2008;39(3):1370–1382. doi:10.1016/j.neuroimage.2007.09.071

24. Owen M, Ingo C, Dewald JPA. Upper Extremity Motor Impairments and Microstructural Changes in Bulbospinal Pathways in Chronic Hemiparetic Stroke. Front Neurol. 2017;8(JUN):1–10. doi:10.3389/fneur.2017.00257

25. McPherson JG, Chen A, Ellis MD, Yao J, Heckman CJ, Dewald JPA. Progressive recruitment of contralesional cortico-reticulospinal pathways drives motor impairment post stroke. J Physiol. 2018;596(7):1211–1225. doi:10.1113/JP274968

26. Takenobu Y, Hayashi T, Moriwaki H, Nagatsuka K, Naritomi H, Fukuyama H. Motor recovery and microstructural change in rubro-spinal tract in subcortical stroke. NeuroImage Clin. 2014;4:201–208. doi:10.1016/j.nicl.2013.12.003

27. Karbasforoushan H, Cohen-Adad J, Dewald JPA. Brainstem and spinal cord MRI identifies altered sensorimotor pathways post-stroke. Nat Commun. 2019;10(1):3524. doi:10.1038/s41467-019-11244-3

28. Ruber T, Schlaug G, Lindenberg R. Compensatory role of the cortico-rubro-spinal tract in motor recovery after stroke. Neurology. 2012;79(6):515–522. doi:10.1212/WNL.0b013e31826356e8

29. Guo J, Liu J, Wang C, et al. Differential involvement of rubral branches in chronic capsular and pontine stroke. NeuroImage Clin. 2019;24(154):102090. doi:10.1016/j.nicl.2019.102090

30. Di Pino G, Pellegrino G, Assenza G, et al. Modulation of brain plasticity in stroke: A novel model for neurorehabilitation. Nat Rev Neurol. 2014;10(10):597–608. doi:10.1038/nrneurol.2014.162

31. Wedeen VJ, Hagmann P, Tseng WYI, Reese TG, Weisskoff RM. Mapping complex tissue architecture with diffusion spectrum magnetic resonance imaging. Magn Reson Med. 2005;54(6):1377–1386. doi:10.1002/mrm.20642

32. Tan ET, Marinelli L, Sperl JI, Menzel MI, Hardy CJ. Multi-directional anisotropy from diffusion orientation distribution functions. J Magn Reson Imaging. 2015;41(3):841–850. doi:10.1002/jmri.24589

33. Demeurisse G, Demol O, Robaye E. Motor Evaluation in Vascular Hemiplegia. Eur Neurol. 1980;19(6):382–389. doi:10.1159/000115178

34. Lyle RC. A performance test for assessment of upper limb function in physical rehabilitation treatment and research. Int J Rehabil Res. 1981;4(4):483–492.

35. Cieslak M, Cook PA, He X, et al. QSIPrep: an integrative platform for preprocessing and reconstructing diffusion MRI data. Nat Methods. 2021;18(7):775–778. doi:10.1038/s41592-021-01185-5

36. Gorgolewski K, Burns CD, Madison C, et al. Nipype: A Flexible, Lightweight and Extensible Neuroimaging Data Processing Framework in Python. Front Neuroinform. 2011;5(August). doi:10.3389/fninf.2011.00013

37. Tustison NJ, Avants BB, Cook PA, et al. N4ITK: Improved N3 Bias Correction. IEEE Trans Med Imaging. 2010;29(6):1310–1320. doi:10.1109/TMI.2010.2046908

38. Fonov V, Evans A, McKinstry R, Almli C, Collins D. Unbiased nonlinear average age-appropriate brain templates from birth to adulthood. Neuroimage. 2009;47:S102. doi:10.1016/S1053-8119(09)70884-5

39. Avants BB, Epstein CL, Grossman M, Gee JC. Symmetric diffeomorphic image registration with cross-correlation: evaluating automated labeling of elderly and neurodegenerative brain. Med Image Anal. 2008;12(1):26–41. doi:10.1016/j.media.2007.06.004

40. Zhang Y, Brady M, Smith S. Segmentation of brain MR images through a hidden Markov random field model and the expectation-maximization algorithm. IEEE Trans Med Imaging. 2001;20(1):45–57. doi:10.1109/42.906424

41. Veraart J, Novikov DS, Christiaens D, Ades-aron B, Sijbers J, Fieremans E. Denoising of diffusion MRI using random matrix theory. Neuroimage. 2016;142(1):394–406. doi:10.1016/j.neuroimage.2016.08.016

42. Merlet SL, Deriche R. Continuous diffusion signal, EAP and ODF estimation via Compressive Sensing in diffusion MRI. Med Image Anal. 2013;17(5):556–572. doi:10.1016/j.media.2013.02.010

43. Yeh FC, Wedeen VJ, Tseng WYI. Generalized q-Sampling Imaging. IEEE Trans Med Imaging. 2010;29(9):1626–1635. doi:10.1109/TMI.2010.2045126

44. Yeh FC, Panesar S, Fernandes D, et al. Population-averaged atlas of the macroscale human structural connectome and its network topology. Neuroimage. 2018;178(April):57–68. doi:10.1016/j.neuroimage.2018.05.027

45. Stinear CM, Barber PA, Smale PR, Coxon JP, Fleming MK, Byblow WD. Functional potential in chronic stroke patients depends on corticospinal tract integrity. Brain. 2007;130(Pt 1):170-180. doi:10.1093/brain/awl333

46. Guggisberg AG, Nicolo P, Cohen LG, Schnider A, Buch ER. Longitudinal Structural and Functional Differences Between Proportional and Poor Motor Recovery After Stroke. Neurorehabil Neural Repair. 2017;31(12):1029–1041. doi:10.1177/1545968317740634

47. Choudhury S, Shobhana A, Singh R, et al. The Relationship Between Enhanced Reticulospinal Outflow and Upper Limb Function in Chronic Stroke Patients. Neurorehabil Neural Repair. 2019;33(5):375–383. doi:10.1177/1545968319836233

48. Glover IS, Baker SN. Both Corticospinal and Reticulospinal Tracts Control Force of Contraction. J Neurosci. 2022;42(15):3150–3164. doi:10.1523/JNEUROSCI.0627-21.2022

49. Krakauer JW, Carmichael ST. Broken Movement. The MIT Press; 2017. doi:10.7551/mitpress/9310.001.0001

50. Bohannon RW. Motricity Index Scores are Valid Indicators of Paretic Upper Extremity Strength Following Stroke. J Phys Ther Sci. 1999;11(2):59–61. doi:10.1589/jpts.11.59

51. Sunderland A, Tinson D, Bradley L, Hewer RL. Arm function after stroke. An evaluation of grip strength as a measure of recovery and a prognostic indicator. J Neurol Neurosurg Psychiatry. 1989;52(11):1267–1272. doi:10.1136/jnnp.52.11.1267

52. Keizer K, Kuypers HGJM. Distribution of corticospinal neurons with collaterals to the lower brain stem reticular formation in monkey (Macaca fascicularis). Exp Brain Res. 1989;74(2):311–318. doi:10.1007/BF00248864

53. Sakai ST, Davidson AG, Buford JA. Reticulospinal neurons in the pontomedullary reticular formation of the monkey (Macaca fascicularis). Neuroscience. 2009;163(4):1158–1170. doi:10.1016/j.neuroscience.2009.07.036

54. Snell RS. Clinical Neuroanatomy. 7th ed. Lippincott Williams & Wilkins; 2010.

55. Jankowska E, Edgley SA. How Can Corticospinal Tract Neurons Contribute to Ipsilateral Movements? A Question With Implications for Recovery of Motor Functions. Neurosci. 2006;12(1):67–79. doi:10.1177/1073858405283392

56. Siegel JM, McGinty DJ. Pontine Reticular Formation Neurons: Relationship of Discharge to Motor Activity. Science (80-). 1977;196(4290):678–680. doi:10.1126/science.193185

57. Schulz R, Frey BM, Koch P, et al. Cortico-Cerebellar Structural Connectivity Is Related to Residual Motor Output in Chronic Stroke. Cereb Cortex. 2017;27(1):635–645. doi:10.1093/cercor/bhv251

58. Guder S, Frey BM, Backhaus W, et al. The Influence of Cortico-Cerebellar Structural Connectivity on Cortical Excitability in Chronic Stroke. Cereb Cortex. 2020;30(3):1330–1344. doi:10.1093/cercor/bhz169

59. Grefkes C, Ward NS. Cortical Reorganization After Stroke. Neurosci. 2014;20(1):56–70. doi:10.1177/1073858413491147

60. Volz LJ, Sarfeld AS, Diekhoff S, et al. Motor cortex excitability and connectivity in chronic stroke: a multimodal model of functional reorganization. Brain Struct Funct. 2015;220(2):1093–1107. doi:10.1007/s00429-013-0702-8

61. Volz LJ, Rehme AK, Michely J, et al. Shaping Early Reorganization of Neural Networks Promotes Motor Function after Stroke. Cereb Cortex. 2016;26(6):2882–2894. doi:10.1093/cercor/bhw034

62. Park C hyun H, Chang WH, Ohn SH, et al. Longitudinal Changes of Resting-State Functional Connectivity During Motor Recovery After Stroke. Stroke. 2011;42(5):1357–1362. doi:10.1161/STROKEAHA.110.596155

63. Carter AR, Astafiev S V., Lang CE, et al. Resting state inter-hemispheric fMRI connectivity predicts performance after stroke. Ann Neurol. 2009;67(3):365–375. doi:10.1002/ana.21905

64. Paul T, Hensel L, Rehme AK, et al. Early motor network connectivity after stroke: An interplay of general reorganization and state-specific compensation. Hum Brain Mapp. 2021;42(16):5230–5243. doi:10.1002/hbm.25612

