## Supplement for "The role of corticospinal and extrapyramidal pathways in motor impairment after stroke"

***Supplementary Table 1:*** *Patient demographics. The sample contains 12 fully recovered and 13 non-fully recovered patients as determined by the ARAT-score (fully recovered = 57 points, non-fully recovered < 57 points).*

| *subject* | *sex* | *affected hemisphere* | *ARAT* | *MI arm* | *MI leg* |
|---|---|---|---|---|---|
| *1* | m | l | 19 | 65 | 59 |
| *2* | f | r | 57 | 83 | 83 |
| *3* | m | r | 38 | 76 | 75 |
| *4* | f | r | 56 | 91 | 75 |
| *5* | m | l | 57 | 99 | 91 |
| *6* | m | l | 35 | 92 | 99 |
| *7* | f | l | 32 | 77 | 83 |
| *8* | f | r | 57 | 76 | 75 |
| *9* | m | r | 49 | 91 | 99 |
| *10* | f | l | 56 | 76 | 75 |
| *11* | m | r | 57 | 91 | 99 |
| *12* | m | l | 57 | 99 | 99 |
| *13* | m | l | 55 | 92 | 91 |
| *14* | m | r | 57 | 99 | 99 |
| *15* | m | r | 57 | 99 | 99 |
| *16* | m | r | 53 | 99 | 99 |
| *17* | m | l | 55 | 92 | 99 |
| *18* | m | l | 44 | 83 | 75 |
| *19* | m | r | 37 | 84 | 75 |
| *20* | m | l | 57 | 99 | 99 |
| *21* | m | l | 57 | 99 | 99 |
| *22* | m | l | 0 | 34 | 34 |
| *23* | m | r | 57 | 99 | 71 |
| *24* | m | l | 57 | 99 | 99 |
| *25* | m | r | 57 | 99 | 99 |

***Supplementary Table 2:*** *Linear regression results when combining gFA derived from one-directional voxels of the ipsilesional CST with gFA derived from two-directional extrapyramidal voxels. The results suggest that ipsilesional CST and extrapyramidal tracts were largely independent with respect to the explanation of behavioral variance in motor impairment. (il = ipsilesional, cl = contralesional, CST = corticospinal tract, reticuloST = reticulospinal tract, rubroST = rubrospinal tract)*

| **DV** | **predictor 1** | **$R^2$** | **p** | **predictor 2** | **$R^2$** | **p** |
|---|---|---|---|---|---|---|
| MI-arm | il CST | 31.94% | 0.003 | cl rubroSt | 51.02% | 0.0004 |
| MI-leg | il CST | 17.71% | 0.036 | il reticuloST | 38.86% | 0.0045 |
| MI-leg | il CST | 17.71% | 0.036 | cl reticuloST | 39.61% | 0.0039 |
| MI-leg | il CST | 17.71% | 0.036 | il rubroST | 38.94% | 0.0044 |
| MI-leg | il CST | 17.71% | 0.036 | cl rubroST | 39.85% | 0.0037 |
